# Risk and Outcome of Second primary malignancy in patients with classical Hodgkin lymphoma

**DOI:** 10.1101/2022.08.06.22278488

**Authors:** Fan Wang

**Affiliations:** Department of Hematology, Tongji Hospital, Tongji Medical College, Huazhong University of Science and Technology, Wuhan, Hubei, The People’s Republic of China

**Keywords:** classical Hodgkin Lymphoma, SEER, SIR, second primary malignancies, outcome

## Abstract

**BACKGROUND:** Hodgkin lymphoma survivors demonstrated increased risk of secondary primary malignancies (SPMs), but comprehensive analysis of the risk and outcome of SPMs in classical Hodgkin lymphoma (cHL) patients has not yet been reported.

**METHODS:** Patients with classical Hodgkin Lymphoma from 1975 to 2017 were identified from the Surveillance, Epidemiology and End Results (SEER) database. Standardized incidence ratios (SIRs) were calculated for the risk of solid and hematologic SPMs in cHL patients compared to the general population. The outcome of cHL patients developing SPMs were assessed by performing survival, competing risks regression and cox proportional regression analyses.

**RESULTS:** In a follow-up of 26,493 cHL survivors for 365,156 person years, 3,866 (14.59%) secondary cancers were identified, with an SIR of 2.09 (95% CI: 2.02 - 2.15). The increased risk was still notable after follow-up of 10 years or more, and the risk is more pronounced for patients with female gender, younger age, advanced stage, chemotherapy and radiation therapy. The overall survival is worse for cHL patients with SPMs after 5 years of follow-up (P < 0.0001). The main cause of death for cHL patients with SPMs is not cHL but other causes including SPMs. Multivariate Cox regression analysis confirmed SPMs as an independently adverse prognostic factor for cHL survivors (hazard ratio, 1.08; 95% CI, 1.03-1.14, P□=□0.002).

**CONCLUSIONS:** There is a significantly increased risk of developing SPMs for cHL survivors. The overall survival is worse for cHL patients and SPMs is an independent prognostic factor for cHL.

## Introduction

Classical Hodgkin lymphoma (cHL) is a relatively rare type of malignancy with an incidence of 2-3 cases per 100,000 people per year in western populations^(Yung & Linch 2003)^, but it is one of the more frequent lymphomas that accounts for 15% to 25% of all lymphomas and approximately 95% of Hodgkin lymphoma^(Jaffe et al. 2016)^. Some studies have shown that Epstein-Barr virus infection could be related to 25-40% of cHL cases, however, there are no clearly defined risk factors for the cause and development of cHL^(Urayama et al. 2012)^. The pathologic hallmark of cHL is the presence of the characteristic multinucleated giant Hodgkin and Reed-Sternberg (HRS) cells within the inflammatory including B lymphocytes, T lymphocytes, eosinophils and macrophages^(Greaves et al. 2013)^. Epidemiology research showed that cHL has a bimodal age distribution with a first peak in patients at the age of 20-30 years and a second peak in patients older than 55 years^(Brice et al. ; Eichenauer et al. 2018)^. Classical Hodgkin lymphomas are histologically heterogenous and are generally subclassified into four subgroups: lymphocyte-rich (LRCHL), lymphocyte-depleted (LDCHL), nodular sclerosis (NSCHL) and mixed cellularity (MCCHL)^(Campo et al. 2017)^. NSCHL is the most common histological subtype of cHL, comprising up to 70% of all cHL cases and is mostly frequent in young adults with the peak incidence at 15-35 years of age^(Lin et al. 2010)^. LDCHL is the rarest subtype, comprising less than 1% of cHL cases^(Wang et al. 2019)^. MCCHL accounts for about 15-30% of cHL cases and is mostly found in adult patients older than 55 years of age, and it is reported with an association to infection of Epstein-Barr virus^(Connors et al. 2020)^. LRCHL is an uncommon subtype of cHL, making up about 5% of all CHL cases^(Wang et al. 2019)^.

Classical Hodgkin Lymphoma is generally viewed as a highly curable cancer with standard first-line chemotherapy and radiotherapy in some cases^(Myint et al. 2020)^. Advances in therapeutic armamentarium for patients with cHL has significantly increased the cure rates, reaching 90%^(Shanbhag & Ambinder 2018)^. Long-term follow-up of Hodgkin lymphoma survivors demonstrated increased risk of secondary primary malignancies (SPMs), which now stand for one of the most important late morbidity^(Ng et al. 2002a; Ng et al. 2002b; Schaapveld et al. 2015)^. To our knowledge, however, comprehensive analysis of the risk of SPMs in cHL survivors has not yet been reported. Furthermore, there is a lack of information on the outcome of SPMs diagnosed in patients with cHL. In this study, we investigated the risk of SPMs development specifically in cHL patients as compared to the general population using the national Cancer Institute’s Surveillance, Epidemiology and End Results (SEER) database. We also evaluated the SPMs incidence based on age, gender, race, stage and subtype of cHL cases. Furthermore, we explored the impact of SPMs occurrence on the prognosis of cHL patients.

## Materials and Methods

### Data Source

The Surveillance, Epidemiology, and End Results (SEER) Program of the National Cancer Institute (NCI) is a reliable source of cancer incidence and survival data that covers around 35% of the population in the United States. The SEER*Stat software (version 8.3.9.1; NCI, Bethesda, MD, USA) was used to obtain the data. Using the US population-based SEER 9 Registry Custom Data, Nov 2019 Sub (1975-2017), which covers approximately 9.4% of the U.S. population (based on 2010 census) from five states (Connecticut, Hawaii, Iowa, New Mexico, and Utah) and four metropolitan areas (Detroit, Atlanta, San Francisco-Oakland, and Seattle-Puget Sound), cHL patients diagnosed from January 1975 to December 2017 were selected. The cHL cases were identified according to the Lymphoma Subtype Recode/WHO 2008, which is updated for Hematopoietic codes on the basis of the World Health Organization (WHO) International Classification of Diseases for Oncology, 3rd Edition (ICD-O-3), and the WHO Classification of Tumors of Haematopoietic and Lymphoid Tissues (2008). The cHL pathologic subtypes include lymphocyte rich cHL, mixed cellularity cHL, lymphocyte depleted cHL, nodular sclerosing cHL, cHL not otherwise specified (NOS). Cases diagnosed during an autopsy and those who were lost to follow-up were excluded. Cases with only known age and only malignant behavior were selected. SPMs was defined as a metachronous malignancy that developed at least 6 months after diagnosis of cHL. The multiple primary standardized incidence ratio (MP-SIR) session of the SEER*Stat software was used to calculate the SIR of SPMs for cHL survivors. SIR measures the relative risk of two cancers, it was calculated as the ratio of the observed (O) number of second cancer cases in the study group and the expected (E) number of second cancer cases in the general population. Absolute excess risk (AER) is an absolute measure of the clinical burden of the additional cancer occurrence in the study population. It is reported as the number of excess events per 10,000 person per year, and is calculated as: ((Observed count - Expected count) * 10,000) / Person years at risk. Kruskal Wallis and Wilcoxon rank sum tests were used to compare the differences in the time to the development of a SPMs based on the different cancer types. The confidence intervals (CIs) at the 95% level for MP-SIRs were calculated with the exact method. We also calculated MP-SIRs for solid tumors and hematologic malignancies with respect to the latency from the index cHL diagnosis (0.5-1, 1-5, 5-10, or >10 years). Data on patients’ demographic profile including age, gender, race, subtype, stage, types of SPMs, survival status and cause of death were extracted. The factor of age was categorized into three groups: < 20 years old, 20-59 years old and 60+ years old. The stage information for cHL is according to the Ann Arbor staging system, and the stage of “N/A, Unknown, Blank” were excluded, the Ann Arbor Stage I and II were combined as “Stage Early”, and the Ann Arbor Stage III and Stage IV were combined as “Stage Advanced”. Cause-of-death information was taken from the “SEER cause-specific death classification” field in the SEER data. Overall survival time was calculated from date of cHL diagnosis to death or last follow up. The approval of local ethics committee was not needed since all the SEER data used in the present study are publicly available.

### Propensity score matching

To minimize the bias effect of potential confounders on selection bias, propensity score matching (PSM) was carried out by using the R “MatchIt” package v4.1.0 ^(Ho et al. 2011)^. The covariates of age, race, sex, stage and subtype for cHL were incorporated into matching analysis with the ration of 1:2 to balance differences in baseline clinical characteristics between cHL cases with or without SPMs, yielding a group of 4500 cHL subjects with SPMs and a group of 9000 subjects without SPMs. To compare the overall survival between cHL patients with SPMs and without SPMs, the “survival” package (version 3.2.7)^(Therneau 2021)^ in R was used for survival analysis with log-rank tests and the “survminer” package (version 0.4.8)^(Kassambara et al. 2020)^ was used for drawing the Kaplan-Meier survival curve. The cumulative incidence function (CIF) of cancer death from the initial cHL diagnosis was measured with a competing-risk Fine-Gray model, treating death of other causes instead of cHL as a competing event. The CIF analysis was performed by using the “cmprsk” package (version 2.2.10)^(Gray 2020)^ in R, and the differences in incidence across strata were compared with the Gray test^(Scrucca et al. 2007)^. To identify the potential independent risk factors for overall survival of cHL mortality, univariate and multivariate Cox proportional regression analyses were performed by using the “survival” package as well, the hazards ratio (HR) estimates and 95% confidence intervals (95% CIs) were reported.

### Statistical analysis

All statistical analysis of the present study was carried out using the R program language (http://www.r-project.org/, version 4.0.4; R Foundation for Statistical Computing, Vienna, Austria). Comparisons between the two groups of continuous data and categorical data were performed using independent T-test and chi-square test, respectively. The Fisher exact method was applied when the smallest expected value is less than five. All *P* values were two-sided and a *P* value < 0.05 was considered to be statistically significant.

## Results

### Observed risk of SPMs in cHL versus the general population

In total 26,493 patients were diagnosed with cHL as a primary malignancy in the SEER 9 registry, Nov 2019 Sub (1975-2017) from January 1975 to December 2017. Among these patients, 3,866 (14.59%) patients with second cancers were identified, with a SIR of 2.09 (95% CI: 2.02 - 2.15, *P* < 0.05), and an AER of 55.16. There was a significantly higher risk of malignancies in the following sites when compared with the general population: oral cavity and pharynx, digestive system (esophagus, stomach, liver, anus, etc.), respiratory system, bone and joints, soft tissue, skin, breasts, female genital system, and endocrine system. Additionally, leukemia, lymphoma, and mesothelioma occurred more frequently than in the general population (Table 1). Among hematological malignancies, leukemia and lymphoma were significantly increased with an SIR of 4.41 (*P* < 0.05, AER = 4.68) and 4.74 (*P* < 0.05, AER = 9.42), respectively. The risk of developing a non-Hodgkin lymphoma was greater than five times higher in cHL survivors than in the general population (NHL, SIR = 5.35, *P* < 0.05, AER = 9.40). Acute myeloid leukemia (SIR = 10.06, *P* < 0.05, AER = 3.50) and acute lymphocytic leukemia (SIR = 5.21, *P* < 0.05, AER = 0.33) were the most commonly occurring types of leukemia after primary cHL. Additionally, the incidence of chronic myeloid leukemia (SIR = 2.06, *P* < 0.05, AER = 0.21) was significantly increased in patients previously diagnosed with cHL. However, the incidence of chronic lymphocytic leukemia was significantly decreased in cHL survivors than in general population (SIR = 0.55, *P* < 0.05, AER = 0.24). Moreover, no significant increased risk of Kaposi Sarcoma was observed in cHL survivors.

**Table 1.** Risk of second primary malignancies in patients with cHL reported in the SEER database between January 1975 and December 2017.

|  | Observed Cases | Expected Cases | MP-SIR | P value | CI Lower | CI Upper | AER |
| --- | --- | --- | --- | --- | --- | --- | --- |
| All Sites | 3866.00 | 1851.66 | 2.09 | < 0.05 | 2.02 | 2.15 | 55.16 |
| Oral Cavity and Pharynx | 138.00 | 52.15 | 2.65 | < 0.05 | 2.22 | 3.13 | 2.35 |
| Lip | 13.00 | 4.10 | 3.17 | < 0.05 | 1.69 | 5.43 | 0.24 |
| Tongue | 42.00 | 15.11 | 2.78 | < 0.05 | 2.00 | 3.76 | 0.74 |
| Salivary Gland | 37.00 | 5.10 | 7.26 | < 0.05 | 5.11 | 10.01 | 0.87 |
| Digestive System | 552.00 | 318.60 | 1.73 | < 0.05 | 1.59 | 1.88 | 6.39 |
| Esophagus | 47.00 | 19.19 | 2.45 | < 0.05 | 1.80 | 3.26 | 0.76 |
| Stomach | 71.00 | 27.28 | 2.60 | < 0.05 | 2.03 | 3.28 | 1.20 |
| Anus, Anal Canal and Anorectum | 32.00 | 7.19 | 4.45 | < 0.05 | 3.05 | 6.29 | 0.68 |
| Liver, Gallbladder, Intrahep Bile Duct and Other Biliary | 57.00 | 36.53 | 1.56 | < 0.05 | 1.18 | 2.02 | 0.56 |
| Respiratory System | 680.00 | 238.86 | 2.85 | < 0.05 | 2.64 | 3.07 | 12.08 |
| Bones and Joints | 17.00 | 3.38 | 5.03 | < 0.05 | 2.93 | 8.05 | 0.37 |
| Soft Tissue including Heart | 71.00 | 12.19 | 5.82 | < 0.05 | 4.55 | 7.34 | 1.61 |
| Skin excluding Basal and Squamous | 157.00 | 111.12 | 1.41 | < 0.05 | 1.20 | 1.65 | 1.26 |
| Breast | 622.00 | 265.03 | 2.35 | < 0.05 | 2.17 | 2.54 | 9.78 |
| Female Genital System | 138.00 | 102.85 | 1.34 | < 0.05 | 1.13 | 1.59 | 0.96 |
| Male Genital System | 289.00 | 311.50 | 0.93 | 1.30 | 0.82 | 1.04 | 0.62 |
| Urinary System | 197.00 | 142.56 | 1.38 | < 0.05 | 1.20 | 1.59 | 1.49 |
| Endocrine System | 188.00 | 55.27 | 3.40 | < 0.05 | 2.93 | 3.92 | 3.63 |
| Mesothelioma | 15.00 | 3.73 | 4.02 | < 0.05 | 2.25 | 6.63 | 0.31 |
| All Lymphatic and Hematopoietic Diseases | 684.00 | 165.43 | 4.13 | < 0.05 | 3.83 | 4.46 | 14.20 |
| Lymphoma | 436.00 | 91.98 | 4.74 | < 0.05 | 4.31 | 5.21 | 9.42 |
| Hodgkin Lymphoma | 14.00 | 13.10 | 1.07 | 0.83 | 0.58 | 1.79 | 0.02 |
| Non-Hodgkin Lymphoma | 422.00 | 78.89 | 5.35 | < 0.05 | 4.85 | 5.89 | 9.40 |
| Leukemia | 221.00 | 50.06 | 4.41 | < 0.05 | 3.85 | 5.04 | 4.68 |
| Acute Lymphocytic Leukemia | 15.00 | 2.88 | 5.21 | < 0.05 | 2.92 | 8.60 | 0.33 |
| Chronic Lymphocytic Leukemia | 11.00 | 19.92 | 0.55 | < 0.05 | 0.28 | 0.99 | 0.24 |
| Acute Myeloid Leukemia | 142.00 | 14.12 | 10.06 | < 0.05 | 8.47 | 11.85 | 3.50 |
| Chronic Myeloid Leukemia | 15.00 | 7.29 | 2.06 | < 0.05 | 1.15 | 3.39 | 0.21 |
| Kaposi Sarcoma | 14 | 8.21 | 1.7 | 0.06 | 0.93 | 2.86 | 0.16 |

Abbreviations: MP-SIR, multiple primary standardized incidence ratio; CI, confidence interval; AER, absolute excess risk.

### Relative risk of SPMs in cHL patients according to latency period

Exploring the latency of developing SPMs after the diagnosis cHL, the risk compared to the U.S. general population was increased across all latency periods. The risk for all sites was elevated and almost stable from 6 months to 119 months, however, the SIR was obviously increased after 120 months (Table 2, SIR 2.34; 95% CI 2.25 - 2.43, P < 0.05). Furthermore, based on cancer types, the risk of developing second solid tumors was variable within all the follow-up period, and the maximum risk was observed after 120 months (Table 2, SIR 2.26; 95% CI 2.16 - 2.35, *P* < 0.05); the risk of developing the second hematological malignancies was maximum within 12-59 months after diagnosis of cHL (Table 2, SIR 6.26; 95% CI 5.44 - 7.17, *P* < 0.05). The risk for the development of an extranodal cHL was relatively high within 6-11 months (Table 2, SIR 102.13; 95% CI 2.59 - 569.02, *P* < 0.05), however, the risk was not significantly elevated compared to the general population after one year. The risk for the development of non-Hodgkin Lymphoma was increased within 6-11 months and was steadily decreased in the subsequent periods (Table 2), while the risk for the development of Acute Lymphoblastic Leukemia and Acute Myeloid Leukemia were increased within 12-59 months and was steadily decreased after 60 months (Table 2). The significantly elevated risk for the development of Chronic Myeloid Leukemia was only observed within 60-119 months (Table 2, SIR 3.95; 95% CI 1.45 - 8.59, *P* < 0.05).

**Table 2.**
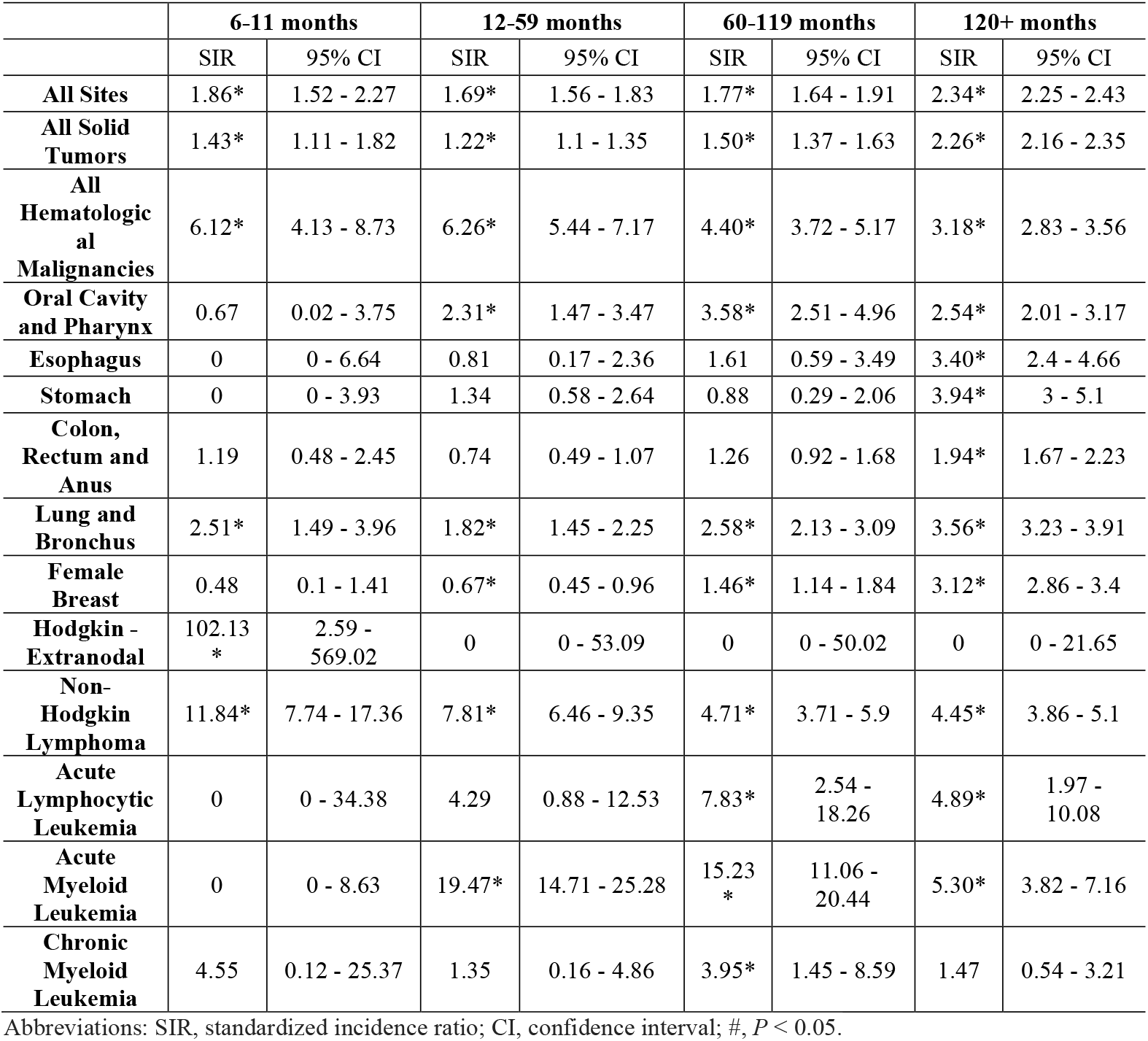
Risk of second primary malignancies in patients with cHL by latency period between January 1975 and December 2017.

### Risk of SPMs by clinical and demographic factors

A further analysis was performed to investigate the relationship of clinical and demographic factors and SPMs. A forest plot of SPMs incidence in cHL patients was shown in Figure 1. Analysis based on gender revealed that the risk of SPMs was higher in women than in men either for aggregate SPMs or for the solid/hematological categories (Figure 1). Additionally, the risk of SPMs in different age groups was explored. For young cHL patients (< 20 years old), the relative risk of aggregate SPMs was quite high (MP-SIR, 5.21; 95% CI, 4.71-5.75; Figure 1A), the risk decreased in patients aged between 20-59 years (MP-SIR, 2.18; 95% CI, 2.10-2.26; Figure 1A) and aged over 60 years (MP-SIR, 1.37; 95% CI, 1.27-1.47; Figure 1A). For the second hematological malignancies, young cHL group showed similar risk with 20-59-year-old group (MP-SIR, 4.59; 95% CI, 3.35-6.15; MP-SIR, 4.29; 95% CI, 3.92-4.69; respectively. Figure 1B), while the risk was slightly decreased in group aged over 60 years (MP-SIR, 3.63; 95% CI, 3.09-4.24; Figure 1B). However, for the second solid tumors, although the risk was still quite high for young cHL patients (MP-SIR, 5.25; 95% CI, 4.71-5.83; Figure 1C), the risk for patients aged over 60 years was only 15% higher than general population (MP-SIR, 1.15; 95% CI, 1.05-1.25; Figure 1C). For the analysis by race, the risk of developing SPMs (especially the secondary hematological malignancies) was higher among “other race (Asians, American Indians, Native Americans, and Pacific Islanders)” as compared to African Americans and the Whites, while African Americans and the Whites showed similar risk of developing SPMs (Figure 1). Based on the stages of cHL, it was noted that the early stage and advanced stage subgroup of cHL patients had the similar risk of developing aggregate SPMs and solid tumors (Figure 1A & C), however, the advanced stage subgroup had comparatively much higher risk of developing a secondary hematological malignancy (MP-SIR, 4.82; 95% CI, 4.20-5.51; Figure 1B). According of the classification cHL, the nodular sclerosis subgroup of cHL patients had the highest risk of developing aggregate SPMs and solid tumors (Figure 1A & C), while the lymphocyte-rich subgroup had the highest risk of developing a secondary hematological malignancy (MP-SIR, 6.99; 95% CI, 5.32-9.01; Figure 1B). Compared to patients without chemotherapy, the chemotherapy group had higher risk to develop secondary hematological malignancies (Figure 1B), but less risk of secondary solid tumors (Figure 1C). Patients with radiation therapy had much higher risk of secondary solid tumors (Figure 1C), but slightly increased risk of secondary hematological malignancies in relative to those without radiation therapy (Figure 1B).

**Figure 1.**
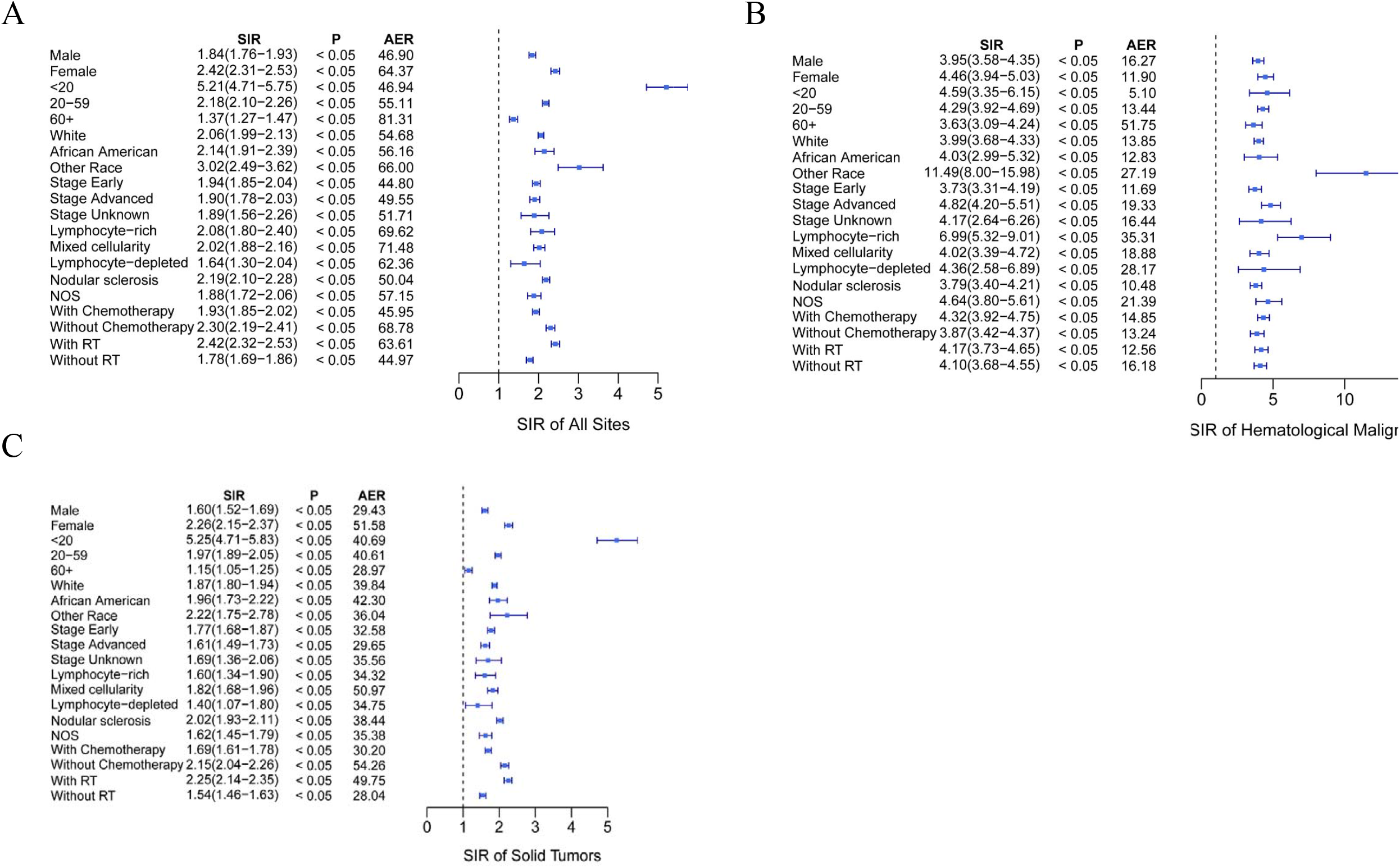
Forest plot of the standardized incidence ratios of secondary malignancies according to clinical and demographic factors in patients with classical Hodgkin Lymphoma from the SEER database between January 1975 and December 2017. (A), (B) and (C) indicates SIR analysis of all sites, hematological malignancies and solid tumors, respectively. *P* < 0.05, compared to the general population. Ann Arbor Stage I and II were combined as Stage Early, while Ann Arbor Stage III and Stage IV were combined as Stage Advanced. RT, radiation therapy; SIR, standardized incidence ratio; cHL, classical Hodgkin Lymphoma; AER, absolute excess risk.

### Outcome of cHL patients developing SPMs

To exclude the bias effect of demographic factors, propensity score matching (PSM) was employed to minimize confounding effects between groups with or without SPMs. The baseline characteristics (age, race, sex, stage, subtype for cHL) were incorporated into matching analysis with the ration of 1:2. For each comparison, all the characteristics were well matched (Table 3). The overall survival was not significantly different between the cHL only group and the group with SPMs before 5 years of follow-up (*P* = 0.55, Figure 2A). However, the cHL with SPMs group showed significantly worse overall survival versus the cHL only group after 5 years of follow-up (*P* < 0.0001; Figure 2B). By applying the competing risk analysis, it is evident that the cumulative incidence of death from cHL is significantly higher in the cHL only group compared to the group with SPMs (*P* < 0.001; Figure 2C), and the cumulative incidence of death from other causes rather than directly from cHL was significantly lower in the cHL only group (*P* < 0.001; Figure 2D).

**Table 3.** Characteristics of cHL Patients with SPMs matched patients without SPMs.

| Characteristics | SPMs |  | P-value |
| --- | --- | --- | --- |
|  | Yes<br>(N=4500) | No<br>(N=9000) |  |
| **Sex** |  |  |  |
| Male | 2301 (51.1%) | 4472 (49.7%) | 0.118 |
|  | Yes<br>(N=4500) | No<br>(N=9000) |  |
| Female | 2199 (48.9%) | 4528 (50.3%) |  |
| **Age** |  |  |  |
| <20 | 231 (5.1%) | 440 (4.9%) | 0.581 |
| 20-59 | 2638 (58.6%) | 5355 (59.5%) |  |
| 60+ | 1631 (36.2%) | 3205 (35.6%) |  |
| **Race** |  |  |  |
| White | 3946 (87.7%) | 7903 (87.8%) | 0.971 |
| African American | 378 (8.4%) | 745 (8.3%) |  |
| Other | 176 (3.9%) | 352 (3.9%) |  |
| **Stages** |  |  |  |
| Early | 2560 (56.9%) | 4949 (55.0%) | 0.106 |
| Advanced | 1717 (38.2%) | 3597 (40.0%) |  |
| Unknown | 223 (5.0%) | 454 (5.0%) |  |
| **Subtype** |  |  |  |
| Lymphocyte-rich | 221 (4.9%) | 417 (4.6%) | 0.732 |
| Mixed cellularity | 840 (18.7%) | 1762 (19.6%) |  |
| Lymphocyte-depleted | 94 (2.1%) | 189 (2.1%) |  |
| Nodular sclerosis | 2445 (54.3%) | 4864 (54.0%) |  |
| NOS | 900 (20.0%) | 1768 (19.6%) |  |
| **Chemotherapy** |  |  |  |
| Yes | 2869 (63.8%) | 5746 (63.8%) | 0.934 |
| No | 1631 (36.2%) | 3254 (36.2%) |  |
| **RT** |  |  |  |
| Yes | 1874 (41.6%) | 3606 (40.1%) | 0.082 |
| No | 2626 (58.4%) | 5394 (59.9%) |  |

\* Ann Arbor Stage I and II were combined as Stage Early, while Ann Arbor Stage III and Stage IV were combined as Stage Advanced.

**Figure 2.**
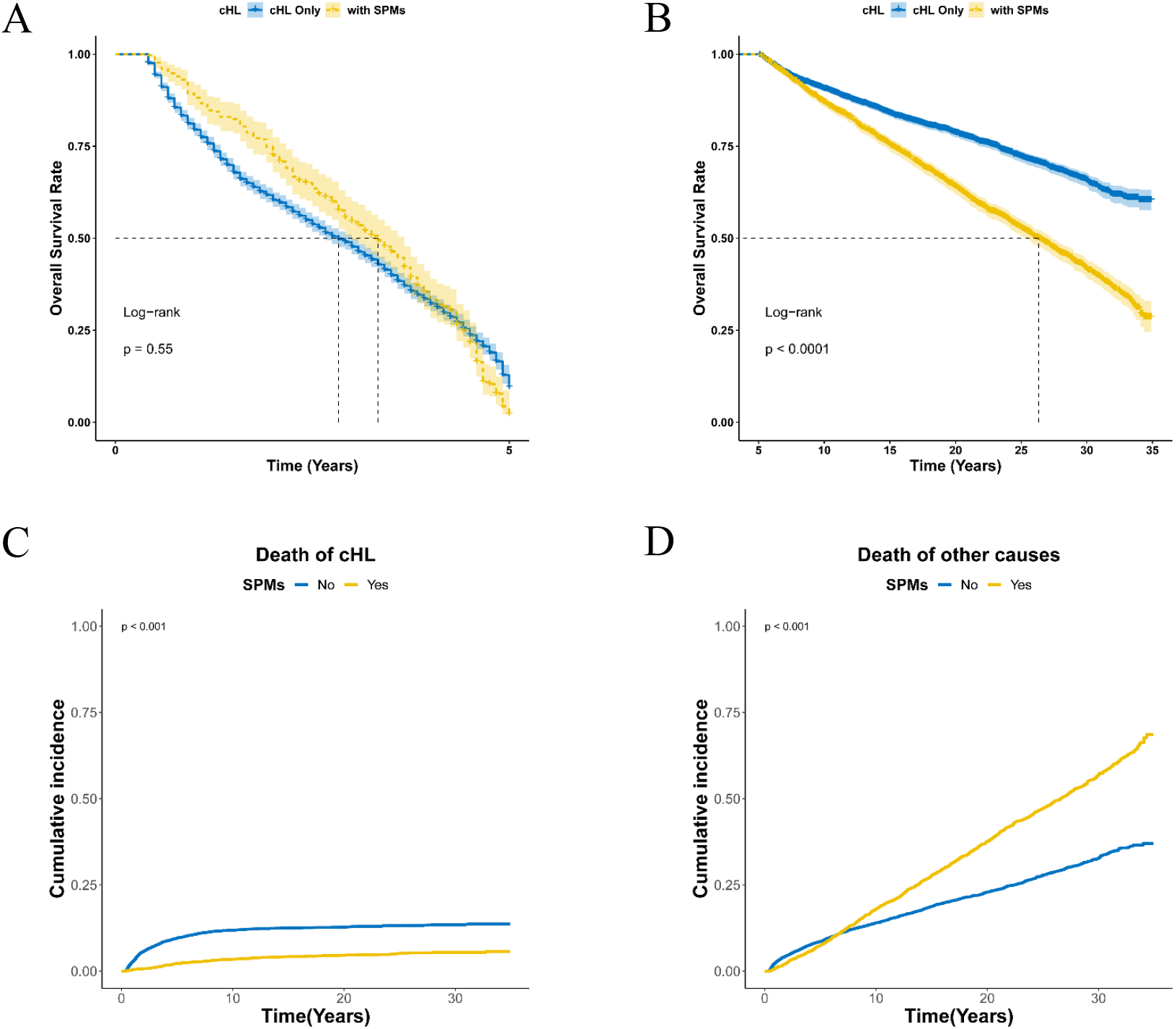
Outcome of cHL patients developing SPMs. Overall survival of cHL patients in cHL only group compared to the group with SPMs before (A) and after (B) 5 years of follow-up was shown, respectively. The cumulative incidence of death from cHL (C) and other causes (D) with the two groups was also shown, respectively. cHL, classical Hodgkin Lymphoma; SPMs, second primary malignancies.

### Cox regression analysis of risk factors for overall survival of cHL patients

Univariate Cox proportional hazard regression analysis for overall survival suggested that older age (*P*□<□0.001, Figure 3A), Male gender (HR 1.51; *P*□<□0.001, Figure 3A), Ethnicity of Asians, American Indians, Native Americans, and Pacific Islanders (HR 1.30; *P*□<□0.001, Figure 3A), subtypes (*P*□<□0.001, Figure 3A), advanced stage (HR 1.81; *P*□<□0.001, Figure 3A), with SPMs (HR 1.15; *P*□<□0.001, Figure 3A) were associated with worse overall survival, radiation therapy instead of chemotherapy was associated with better overall survival (HR 0.46; *P*□<□0.001, Figure 3A). Similar results were obtained with the multivariate logistic regression analysis except little differences: For the variable subtypes, only the subtype of mixed cellularity (HR 1.27; *P*□<□0.001, Figure 3B) and lymphocyte-depleted (HR 2.10; *P*□<□0.001, Figure 3B) other than the subtypes of nodular sclerosis and NOS were independent prognostic factors of cHL overall survival; For the variable race, African-American ethnicity was also associated with worse prognosis (HR 1.18; *P*□<□0.001, Figure 3B); Either chemotherapy or radiation therapy was associated with better prognosis (*P*□<□0.001, Figure 3B).

**Figure 3.**
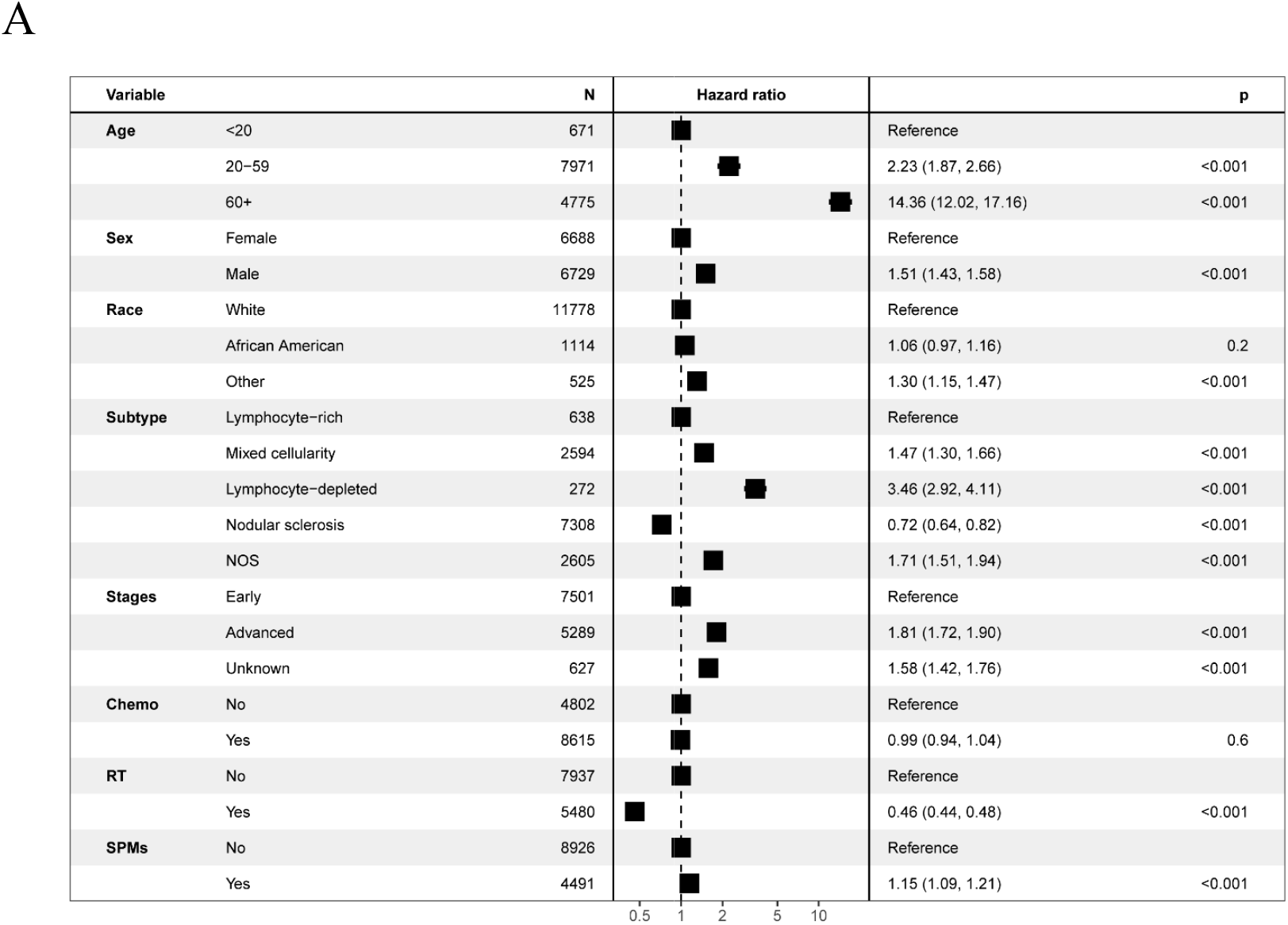
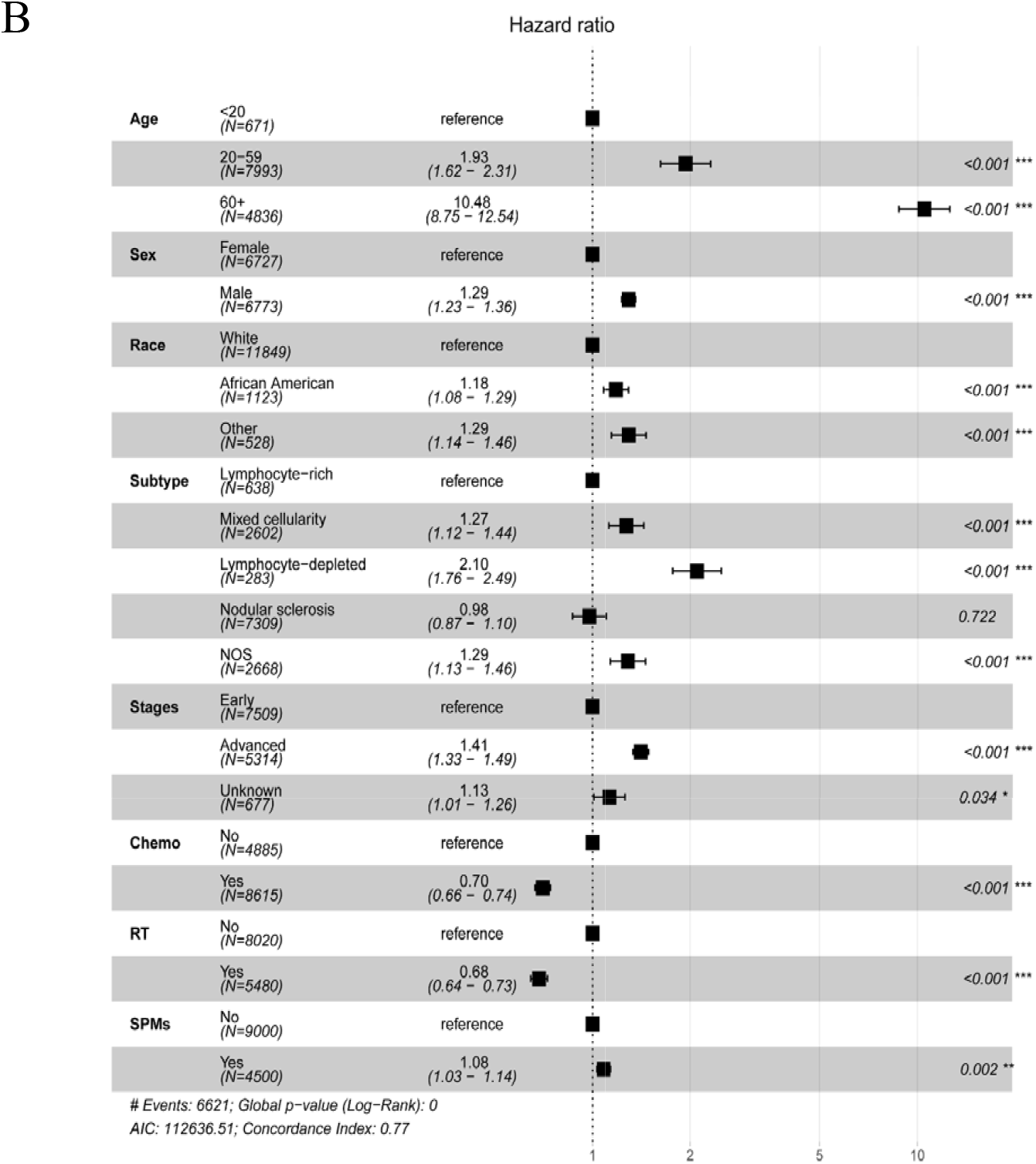
Univariate (A) and multivariate (B) logistic regression analyses for predictors of overall survival in cHL patients after propensity score matching. * Ann Arbor Stage I and II were combined as Stage Early, while Ann Arbor Stage III and Stage IV were combined as Stage Advanced; Race (other): Asians, American Indians, Native Americans, and Pacific Islanders; NOS, not otherwise specified; SPMs, second primary malignancies.

## Discussion

Some studies have shown that Hodgkin Lymphoma patients are at high-risk for developing SPMs^(Schaapveld et al. 2015)^. Nevertheless, systematical studies regarding SPMs among classical Hodgkin Lymphoma patients has not been reported. In the current study, 26,493 patients with cHL as primary malignancies were identified in the SEER 9 registry, Nov 2019 Sub (1975-2017) from January 1975 to December 2017. Among the cHL survivors, we found there was a significantly increased risks of secondary hematological malignancies and solid tumors, such as acute myeloid leukemia, acute lymphocytic leukemia, chronic myeloid leukemia, non-Hodgkin lymphoma, bone and soft tissue tumors, oral cavity and pharynx cancer, digestive system cancer, skin cancer, endocrine tumors, respiratory system cancer and breast cancer, which is similar to some previous studies showed that survivors of Hodgkin Lymphoma had an increased risk of second cancer, but our study has a larger study population and longer span of observation period^(Kumar et al. 2018; Leeuwen et al. 1994; Schaapveld et al. 2015)^. Moreover, increased SIR of SPMs was observed among cHL patients throughout all the latency periods (6-120 months), even after 120 months. Specifically, the risk of developing second solid tumors such as breast cancer was maximum after 120 months, while the risk of developing the second hematological malignancies was maximum within 12-59 months after diagnosis of cHL. The risk for the development of non-Hodgkin Lymphoma was increased within 6-11 months and was steadily decreased thereafter. Thus, it is essential to increase awareness of the incurrence of SPMs during short-term and long-term follow-ups of cHL survivors. To our knowledge, this is the first systematical study about SPMs among classical Hodgkin Lymphoma patients.

In this study, we found female survivors of cHL had higher risk of SPMs, which was also noted by a meta-analysis^(Ibrahim et al. 2012)^. Furthermore, we found that increased risk of SPMs was associated with younger age that may be related with more aggressive and multiple courses of chemotherapy regime applied in the younger patients, since some previous studies have shown chemotherapy is followed by substantial risk of hematological malignancies such as leukemia and NHL^(Swerdlow et al. 2011)^. For the analysis by race, the risk of developing SPMs, especially the secondary hematological malignancies, was highest in “other race (Asians, American Indians, Native Americans, and Pacific Islanders)” in relative to African Americans and the Whites, while African Americans and the Whites showed similar risk of developing SPMs. The disparities may be due to the genetic background and socioeconomic status of different ethnicity group. Compared to patients with early stage of cHL, the advanced stage group demonstrated an increased risk of secondary hematological malignancies instead of secondary solid tumors, this may also be due to the fact that the advanced stage of cHL patients tend to receive escalated chemotherapy regimen with potentially more toxicity, which was supported by the reports that acute myeloid leukemia (AML) or myelodysplastic syndrome (MDS) were more frequently observed in patients treated with the dose escalated chemotherapy regime such as BEACOPP (bleomycin, etoposide, doxorubicin, cyclophosphamide, vincristine, procarbazine, and prednisone)^(Engert et al. 2009)^. Additionally, SPMs risks were strikingly different based on cHL subtypes: the nodular sclerosis subgroup of cHL patients had the highest risk of developing solid tumors, while the lymphocyte-rich subgroup had the highest risk of developing a secondary hematological malignancy. The basis for the differences of SPMs risks in histologic cHL subtypes is unknown but may reflect a nodular sclerosis subtype being more greatly associated with younger age and being apt to undergo more intensified chemotherapy^(Brice et al.)^.

Over past several decades, advances in chemotherapy and the combination of radiation therapy have significantly increased the cure rate of patients with cHL^(Brice et al.)^. However, concerns over the possible late side effects after chemotherapy or radiation therapy, particularly SPMs need to be considered when planning an optimal treatment regime for a given patient. ABVD (adriamycin, bleomycin, vinblastine and dacarbazine) is considered as the standard chemotherapy regimen for HL patients because of its efficacy and lower toxicity^(Rueda Domínguez et al. 2004)^. Some more intensive chemotherapy regimens such as BEACOPP and escalated BEACOPP (eBEACOPP) were developed for HL with unfavorable risk factors^(Tresckow et al. 2012)^ and advanced-stage Hodgkin lymphoma^(Diehl et al. 2003; Kreissl et al. 2021)^, respectively. Studies had shown that ABVD can induce leukemia^(Delwail et al. 2002)^ and BEACOPP increased the incidence secondary leukemia as compared to ABVD^(André et al. 2020)^, which may be associated with the use of alkylating agents and topoisomerase II inhibitors used in the chemotherapy regime^(Eichenauer et al. 2014; Koontz et al. 2013; Leone et al. 2001)^. For solid tumors, chemotherapy alone showed reduced risk of secondary breast cancer^(Swerdlow et al. 2011)^. In the present study, we found that the chemotherapy group had higher risk to develop secondary hematological malignancies, but less risk of secondary solid tumors, which is consistent with previous studies. In a study of cHL survivors treated with radiation therapy, the 5-year survival after development of SPMs was 38%, and the excess risk of SPMs continues to be elevated after 15-20 years of completing therapy without an appearing plateau^14^. We also found patients with radiation therapy had slightly increased risk of secondary hematological malignancies, and higher risk of secondary solid tumors in relative to those without radiation therapy. Thus, survivors of cHL patients treated with radiation therapy represent a high-risk population for secondary solid tumors, optimal screening strategies for those secondary solid tumors should be developed. Nowadays, cHL patients normally received radiation therapy at lower doses and with smaller fields than previous courses, however, its impact on the long-term side-effects such as SPMs still needs further investigation.

In this study, the outcomes of cHL patients who developed SPMs were compared with matched controls, it is interested to find that the overall survival is worse for cHL patients with SPMs as to those without SPMs after 5 years of follow-up. Although worse overall survival among survivors of HL with second primary head and neck cancer^(Chowdhry et al. 2015)^, breast^(Milano et al. 2010)^, and lung cancer^(Milano et al. 2011)^ had previously been described, to our knowledge, the present study is the largest population-based analysis (365,156 patient-years of follow-up) to systematically investigate the outcomes of SPMs for patients with cHL. Moreover, the results of the current study showed that the cumulative incidence of death from cHL is significantly higher in cHL patients without SPMs than those with SPMs, while the cumulative incidence of death from other causes is significantly higher in cHL patients with SPMs than those without SPMs. It indicates that for cHL patients with SPMs, the main risk of death is not cHL itself directly but other causes such as the second primary cancers, diseases of heart, diabetes mellitus, pneumonia and so on, and our finding is well supported by a large retrospective study showed that the main cause of death among young HL patients was SPMs and cardiovascular diseases instead of HL after 20 years of follow-up^(Aleman et al. 2003)^. Furthermore, in this study, multivariate cox regression analysis confirmed SPMs as an independently prognostic factor for cHL survivors, which had not been reported by other studies.

Although the SEER data has the advantage of including a good deal of patients with long period of follow-up covering a comparatively large geographic area, there are still some limitations. Firstly, the SEER registry lacks information including social economic status, carcinogens exposure, family history, alcohol/smoking consumption history, Epstein-Barr virus status, human immunodeficiency virus status, human papillomavirus status, which might potentially affect SPMs risk. Secondly, details concerning therapy for cHL such as immunotherapy (such as PD-1 antibody), chemotherapy regime are not noted in the SEER database, thus the analysis on different treatment regimen is impossible. Additionally, data about radiation dose and fields, and underreporting radiotherapy are also missing, which will attenuate the generalizability of our findings. Thirdly, there are possibilities of under-ascertainment of SPMs risks due to patient migration outside of SEER program areas. Finally, this study is a retrospective study based on SEER database, which is prone to selection bias, recall bias or misclassification bias just as other retrospective cohort studies did.

## Conclusions

In conclusion, our large population-based study indicates that cHL survivors have an increased risk of developing SPMs versus the general population, and the increase in risk is more pronounced for patients with the characteristics of female gender, younger age, advanced stage, chemotherapy and radiation therapy. Additionally, SPMs risks were strikingly different based on cHL subtypes. Moreover, the overall survival is worse for cHL patients with SPMs as to those without SPMs after 5 years of follow-up, the main cause of death for cHL patients with SPMs is other causes including SPMs, and SPMs is an independently prognostic factor for cHL survivors.

These findings suggest that awareness of the increased risk of subsequent SPMs remains crucial for cHL survivors, and ongoing monitoring and management of SPMs during and after therapy for cHL is paramount to improve the survival of cHL patients.

## Data Availability

The data analyzed in this study are from the SEER database (https://seer.cancer.gov/) that are available to the public.

## Abbreviations

cHL,: classical Hodgkin Lymphoma;
SPMs,: second primary malignancies;
SEER,: Surveillance, Epidemiology, and End Results;
MP-SIR,: multiple primary standardized incidence ratio;
CI,: confidence interval;
AER,: absolute excess risk;
PSM,: propensity score matching;
NOS,: not otherwise specified.

## Acknowledgments

The interpretation of the data is the sole responsibility of the author(s). The author(s) acknowledge the efforts of the National Cancer Institute and the Surveillance, Epidemiology, and End Results (SEER) Program tumor registries in the creation of the SEER database.

## References

Surveillance Research Program, National Cancer Institute SEER*Stat software (seer.cancer.gov/seerstat) version 8.3.9.

Surveillance, Epidemiology, and End Results (SEER) Program (https://www.seer.cancer.gov) SEER*Stat Database: Incidence - SEER Research Data, 9 Registries, mNov 2019 Sub (1975-2017) - Linked To County Attributes - Time Dependent (1990-2017) Income/Rurality, 1969-2018 Counties, National Cancer Institute, DCCPS, Surveillance Research Program, released April 2020, based on the November 2019 submission.

Aleman BM, van den Belt-Dusebout AW, Klokman WJ, Van’t Veer MB, Bartelink H, and van Leeuwen FE. 2003. Long-term cause-specific mortality of patients treated for Hodgkin’s disease. Journal of clinical oncology : official journal of the American Society of Clinical Oncology 21:3431–3439. 10.1200/jco.2003.07.131

André MPE, Carde P, Viviani S, Bellei M, Fortpied C, Hutchings M, Gianni AM, Brice P, Casasnovas O, Gobbi PG, Zinzani PL, Dupuis J, Iannitto E, Rambaldi A, Brière J, Clément-Filliatre L, Heczko M, Valagussa P, Douxfils J, Depaus J, Federico M, and Mounier N. 2020. Long-term overall survival and toxicities of ABVD vs BEACOPP in advanced Hodgkin lymphoma: A pooled analysis of four randomized trials. Cancer Medicine 9:6565–6575. https://doi.org/10.1002/cam4.3298

Brice P, de Kerviler E, and Friedberg JW. Classical Hodgkin lymphoma. The Lancet. 10.1016/S0140-6736(20)32207-8

Campo E, Harris NL, Jaffe ES, Pileri SA, Stein H, and Thiele J. 2017. WHO Classification of Tumours of Haematopoietic and Lymphoid Tissues. Lyon, France: International Agency for Research on Cancer.

Chowdhry AK, McHugh C, Fung C, Dhakal S, Constine LS, and Milano MT. 2015. Second primary head and neck cancer after Hodgkin lymphoma: A population-based study of 44,879 survivors of Hodgkin lymphoma. Cancer 121:1436–1445. https://doi.org/10.1002/cncr.29231

Connors JM, Cozen W, Steidl C, Carbone A, Hoppe RT, Flechtner H-H, and Bartlett NL. 2020. Hodgkin lymphoma. Nature Reviews Disease Primers 6:61. 10.1038/s41572-020-0189-6

Delwail V, Jais J-P, Colonna P, and Andrieu J-M. 2002. Fifteen-year secondary leukaemia risk observed in 761 patients with Hodgkin’s disease prospectively treated by MOPP or ABVD chemotherapy plus high-dose irradiation. British Journal of Haematology 118:189–194. https://doi.org/10.1046/j.1365-2141.2002.03564.x

Diehl V, Franklin J, Pfreundschuh M, Lathan B, Paulus U, Hasenclever D, Tesch H, Herrmann R, Dörken B, Müller-Hermelink H-K, Dühmke E, and Loeffler M. 2003. Standard and Increased-Dose BEACOPP Chemotherapy Compared with COPP-ABVD for Advanced Hodgkin’s Disease. New England Journal of Medicine 348:2386–2395. 10.1056/NEJMoa022473

Eichenauer DA, Aleman BMP, André M, Federico M, Hutchings M, Illidge T, Engert A, and Ladetto M. 2018. Hodgkin lymphoma: ESMO Clinical Practice Guidelines for diagnosis, treatment and follow-up. Annals of Oncology 29:iv19–iv29. 10.1093/annonc/mdy080

Eichenauer DA, Thielen I, Haverkamp H, Franklin J, Behringer K, Halbsguth T, Klimm B, Diehl V, Sasse S, Rothe A, Fuchs M, Böll B, von Tresckow B, Borchmann P, and Engert A. 2014. Therapy-related acute myeloid leukemia and myelodysplastic syndromes in patients with Hodgkin lymphoma: a report from the German Hodgkin Study Group. Blood 123:1658–1664. 10.1182/blood-2013-07-512657

Engert A, Diehl V, Franklin J, Lohri A, Dörken B, Ludwig W-D, Koch P, Hänel M, Pfreundschuh M, Wilhelm M, Trümper L, Aulitzky W-E, Bentz M, Rummel M, Sezer O, Müller-Hermelink H-K, Hasenclever D, and Löffler M. 2009. Escalated-Dose BEACOPP in the Treatment of Patients With Advanced-Stage Hodgkin’s Lymphoma: 10 Years of Follow-Up of the GHSG HD9 Study. Journal of Clinical Oncology 27:4548–4554. 10.1200/jco.2008.19.8820

Gray B. 2020. cmprsk: Subdistribution analysis of competing risks.

Greaves P, Clear A, Coutinho R, Wilson A, Matthews J, Owen A, Shanyinde M, Lister TA, Calaminici M, and Gribben JG. 2013. Expression of FOXP3, CD68, and CD20 at Diagnosis in the Microenvironment of Classical Hodgkin Lymphoma Is Predictive of Outcome. Journal of Clinical Oncology 31:256–262. 10.1200/jco.2011.39.9881

Ho D, Imai K, King G, and Stuart EA. 2011. MatchIt: Nonparametric Preprocessing for Parametric Causal Inference. Journal of Statistical Software 42:28. 10.18637/jss.v042.i08

Ibrahim EM, Abouelkhair KM, Kazkaz GA, Elmasri OA, and Al-Foheidi M. 2012. Risk of second breast cancer in female Hodgkin’s lymphoma survivors: a meta-analysis. BMC Cancer 12:197. 10.1186/1471-2407-12-197

Jaffe ES, Arber DA, Campo E, Quintanilla-Fend L, and Harris NL. 2016. Hematopathology. Philadelphia, PA: Elsevier Health Sciences.

Kassambara A, Kosinski M, and Biecek P. 2020. survminer: Drawing survival curves using ‘ggplot2’.

Koontz MZ, Horning SJ, Balise R, Greenberg PL, Rosenberg SA, Hoppe RT, and Advani RH. 2013. Risk of therapy-related secondary leukemia in Hodgkin lymphoma: the Stanford University experience over three generations of clinical trials. Journal of clinical oncology : official journal of the American Society of Clinical Oncology 31:592–598. 10.1200/JCO.2012.44.5791

Kreissl S, Goergen H, Buehnen I, Kobe C, Moccia A, Greil R, Eichenauer DA, Zijlstra JM, Markova J, Meissner J, Feuring-Buske M, Soekler M, Beck H-J, Willenbacher W, Ludwig W-D, Pabst T, Topp MS, Hitz F, Bentz M, Keller UB, Kühnhardt D, Ostermann H, Hertenstein B, Aulitzky W, Maschmeyer G, Vieler T, Eich H, Baues C, Stein H, Fuchs M, Diehl V, Dietlein M, Engert A, and Borchmann P. 2021. PET-guided eBEACOPP treatment of advanced-stage Hodgkin lymphoma (HD18): follow-up analysis of an international, open-label, randomised, phase 3 trial. The Lancet Haematology 8:e398–e409. 10.1016/S2352-3026(21)00101-0

Kumar V, Garg M, Chandra AB, Mayorga VS, Ahmed S, and Ailawadhi S. 2018. Trends in the Risks of Secondary Cancers in Patients With Hodgkin Lymphoma. Clinical Lymphoma, Myeloma and Leukemia 18:576-589.e571. 10.1016/j.clml.2018.05.021

Leeuwen FEv, Klokman WJ, Hagenbeek A, Noyon R, Belt-Dusebout AWvd, Kerkhoff EHv, Heerde Pv, and Somers R. 1994. Second cancer risk following Hodgkin’s disease: a 20-year follow-up study. Journal of Clinical Oncology 12:312–325. 10.1200/jco.1994.12.2.312

Leone G, Voso MT, Sica S, Morosetti R, and Pagano L. 2001. Therapy Related Leukemias: Susceptibility, Prevention and Treatment. Leukemia & Lymphoma 41:255–276. 10.3109/10428190109057981

Lin R, Jones D, and Ibrahim S. 2010. Hodgkin Lymphoma. In: Jones D, ed. Neoplastic Hematopathology: Experimental and Clinical Approaches. Totowa, NJ: Humana Press, 349–366.

Milano MT, Li H, Constine LS, and Travis LB. 2011. Survival after second primary lung cancer: a population-based study of 187 Hodgkin lymphoma patients. Cancer 117:5538–5547. 10.1002/cncr.26257

Milano MT, Li H, Gail MH, Constine LS, and Travis LB. 2010. Long-term survival among patients with Hodgkin’s lymphoma who developed breast cancer: a population-based study. Journal of clinical oncology : official journal of the American Society of Clinical Oncology 28:5088–5096. 10.1200/jco.2010.29.5683

Myint ZW, Shrestha R, Siddiqui S, Slone S, Huang B, Ramlal R, Monohan GP, Hildebrandt GC, and Saeed H. 2020. Ten-year survival outcomes for patients with early stage classical Hodgkin lymphoma: An analysis from Kentucky Cancer Registry. Hematology/Oncology and Stem Cell Therapy 13:17–22. https://doi.org/10.1016/j.hemonc.2019.08.009

Ng AK, Bernardo MP, Weller E, Backstrand KH, Silver B, Marcus KC, Tarbell NJ, Friedberg J, Canellos GP, and Mauch PM. 2002a. Long-Term Survival and Competing Causes of Death in Patients With Early-Stage Hodgkin’s Disease Treated at Age 50 or Younger. Journal of Clinical Oncology 20:2101–2108. 10.1200/jco.2002.08.021

Ng AK, Bernardo MVP, Weller E, Backstrand K, Silver B, Marcus KC, Tarbell NJ, Stevenson MA, Friedberg JW, and Mauch PM. 2002b. Second malignancy after Hodgkin disease treated with radiation therapy with or without chemotherapy: long-term risks and risk factors. Blood 100:1989–1996. 10.1182/blood-2002-02-0634

Rueda Domínguez A, Márquez A, Gumá J, Llanos M, Herrero J, de las Nieves MA, Miramón J, and Alba E. 2004. Treatment of stage I and II Hodgkin’s lymphoma with ABVD chemotherapy: results after 7 years of a prospective study. Annals of Oncology 15:1798–1804. 10.1093/annonc/mdh465

Schaapveld M, Aleman BMP, van Eggermond AM, Janus CPM, Krol ADG, van der Maazen RWM, Roesink J, Raemaekers JMM, de Boer JP, Zijlstra JM, van Imhoff GW, Petersen EJ, Poortmans PMP, Beijert M, Lybeert ML, Mulder I, Visser O, Louwman MWJ, Krul IM, Lugtenburg PJ, and van Leeuwen FE. 2015. Second Cancer Risk Up to 40 Years after Treatment for Hodgkin’s Lymphoma. New England Journal of Medicine 373:2499–2511. 10.1056/NEJMoa1505949

Scrucca L, Santucci A, and Aversa F. 2007. Competing risk analysis using R: an easy guide for clinicians. Bone Marrow Transplantation 40:381–387. 10.1038/sj.bmt.1705727

Shanbhag S, and Ambinder RF. 2018. Hodgkin lymphoma: A review and update on recent progress. CA: a cancer journal for clinicians 68:116–132. 10.3322/caac.21438

Swerdlow AJ, Higgins CD, Smith P, Cunningham D, Hancock BW, Horwich A, Hoskin PJ, Lister TA, Radford JA, Rohatiner AZS, and Linch DC. 2011. Second Cancer Risk After Chemotherapy for Hodgkin’s Lymphoma: A Collaborative British Cohort Study. Journal of Clinical Oncology 29:4096–4104. 10.1200/jco.2011.34.8268

Therneau TM. 2021. A package for survival analysis in r.

Tresckow Bv, Plütschow A, Fuchs M, Klimm B, Markova J, Lohri A, Kral Z, Greil R, Topp MS, Meissner J, Zijlstra JM, Soekler M, Stein H, Eich HT, Mueller RP, Diehl V, Borchmann P, and Engert A. 2012. Dose-Intensification in Early Unfavorable Hodgkin’s Lymphoma: Final Analysis of the German Hodgkin Study Group HD14 Trial. Journal of Clinical Oncology 30:907–913. 10.1200/jco.2011.38.5807

Urayama KY, Jarrett RF, Hjalgrim H, Diepstra A, Kamatani Y, Chabrier A, Gaborieau V, Boland A, Nieters A, Becker N, Foretova L, Benavente Y, Maynadié M, Staines A, Shield L, Lake A, Montgomery D, Taylor M, Smedby KE, Amini R-M, Adami H-O, Glimelius B, Feenstra B, Nolte IM, Visser L, van Imhoff GW, Lightfoot T, Cocco P, Kiemeney L, Vermeulen SH, Holcatova I, Vatten L, Macfarlane GJ, Thomson P, Conway DI, Benhamou S, Agudo A, Healy CM, Overvad K, Tjønneland A, Melin B, Canzian F, Khaw K-T, Travis RC, Peeters PHM, González CA, Quirós JR, Sánchez M-J, Huerta JM, Ardanaz E, Dorronsoro M, Clavel-Chapelon F, Bueno-de-Mesquita HB, Riboli E, Roman E, Boffetta P, de Sanjosé S, Zelenika D, Melbye M, van den Berg A, Lathrop M, Brennan P, and McKay JD. 2012. Genome-Wide Association Study of Classical Hodgkin Lymphoma and Epstein– Barr Virus Status–Defined Subgroups. JNCI: Journal of the National Cancer Institute 104:240–253. 10.1093/jnci/djr516

Wang H-W, Balakrishna JP, Pittaluga S, and Jaffe ES. 2019. Diagnosis of Hodgkin lymphoma in the modern era. British Journal of Haematology 184:45–59. https://doi.org/10.1111/bjh.15614

Yung L, and Linch D. 2003. Hodgkin’s lymphoma. The Lancet 361:943–951. 10.1016/S0140-6736(03)12777-8

